# Early analysis of the Australian COVID-19 epidemic

**DOI:** 10.1101/2020.04.25.20080127

**Authors:** David J Price, Freya M Shearer, Michael T Meehan, Emma McBryde, Robert Moss, Nick Golding, Eamon J Conway, Peter Dawson, Deborah Cromer, James Wood, Sam Abbott, Jodie McVernon, James M McCaw

## Abstract

As of 18 April 2020, there had been 6,533 confirmed cases of COVID-19 in Australia [1]. Of these, 67 had died from the disease. The daily count of new confirmed cases was declining. This suggests that the collective actions of the Australian public and government authorities in response to COVID-19 were sufficiently early and assiduous to avert a public health crisis — for now. Analysing factors, such as the intensity and timing public health interventions, that contribute to individual country experiences of COVID-19 will assist in the next stage of response planning globally. Using data from the Australian national COVID-19 database, we describe how the epidemic and public health response unfolded in Australia up to 13 April 2020. We estimate that the effective reproduction number was likely below 1 (the threshold value for control) in each Australian state since mid-March and forecast that hospital ward and intensive care unit occupancy will remain below capacity thresholds over the next two weeks.

## Main text

A small cluster of cases of the disease now known as COVID-19 was first reported on December 29, 2019, in the Chinese city of Wuhan [2]. By mid-April 2020, the disease had spread to all global regions, and overwhelmed some the world’s most developed health systems. More than 2 million cases and 130,000 deaths had been confirmed globally, and the vast majority of countries with confirmed cases were reporting escalating transmission [3].

As of 18 April 2020, there were 6,533 confirmed cases of COVID-19 in Australia. Of these, 67 had died from the disease. Encouragingly, the daily count of new confirmed cases had been declining since late March 2020. This suggests that Australia has avoided a “worst-case” scenario — one where planning models estimated a peak daily demand for 35,000 ICU beds by around May 2020, far exceeding the health system’s capacity of around 2,200 ICU beds [4].

The first wave of COVID-19 epidemics, and the government and public responses to them, have varied vastly across the globe. For example, many European countries and the United States are in the midst of explosive outbreaks with overwhelmed health systems [5, 6]. Meanwhile, countries such as Singapore and South Korea had early success in containing the spread, partly attributed to their extensive surveillance efforts and case targeted interventions [7, 8]. However, despite those early successes, Singapore has recently taken additional steps to further limit transmission in the face of increasing importations and community spread [9]. Other locations in the region, including Taiwan, Hong Kong and New Zealand, have had similar epidemic experiences, achieving control through a combination of border, case targeted and social distancing measures.

Analysing key epidemiological and response factors — such as the intensity and timing of public health interventions — that contribute to individual country experiences of COVID-19 will assist in the next stage of response planning globally.

Here we describe the course of the COVID-19 epidemic and public health response in Australia from 22 January up to 13 April 2020 (summarised in Figure 1). We then quantify the impact of the public health response on disease transmission (Figure 2) and forecast the short-term health system demand from COVID-19 patients (Figure 3).

**Figure 1:**
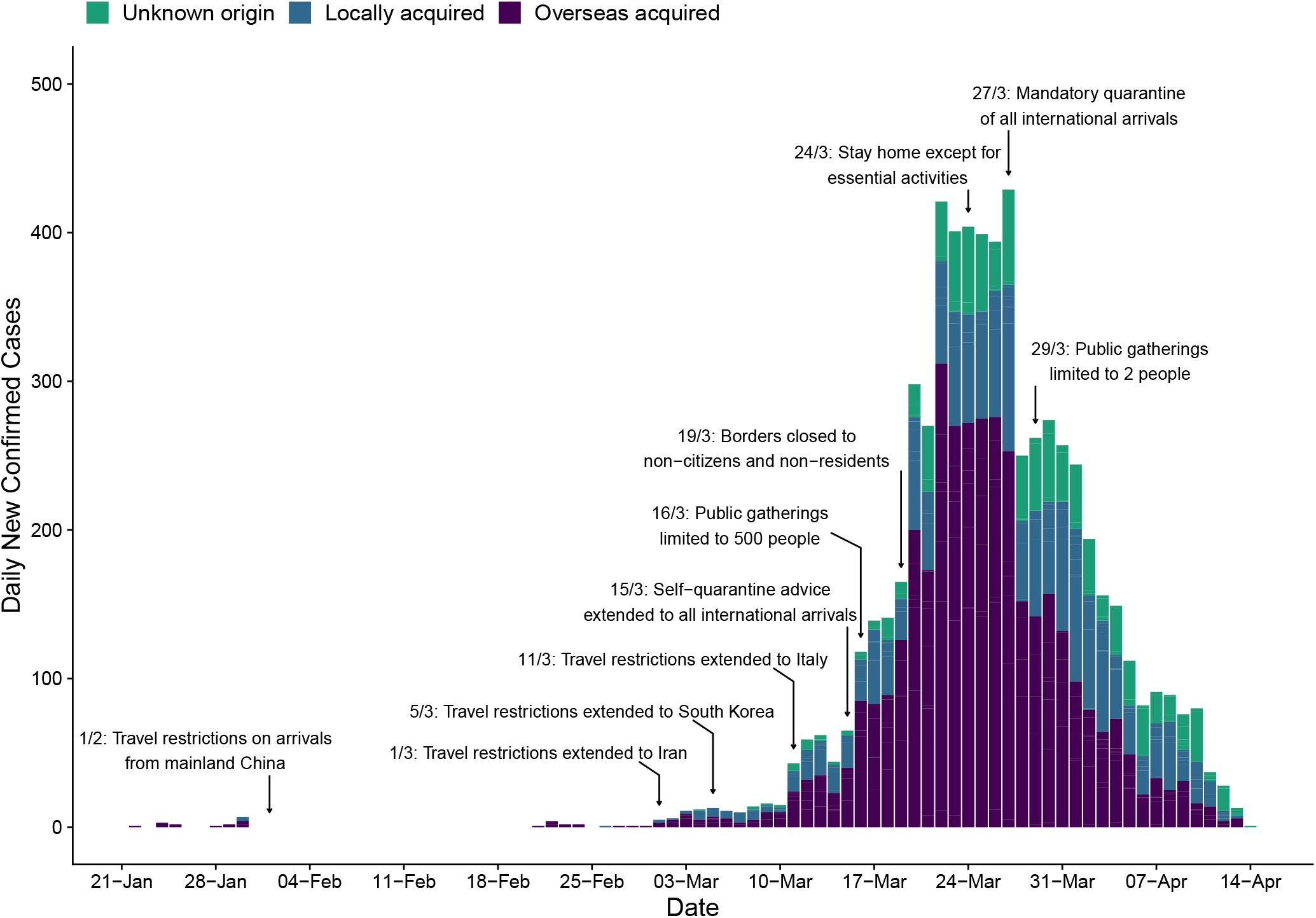
Time series of new daily confirmed cases of COVID-19 in Australia by import status (purple = overseas acquired, blue = locally acquired, green = unknown origin) from 22 January 2020 (first case detected) to 13 April 2020. Dates of selected key border and social distancing measures implemented by Australian authorities are indicated by annotations above the plotted case counts. These measures were in addition to case targeted interventions (case isolation and contact quarantine) and further border measures, including enhanced testing and provision of advice, on arrivals from other selected countries, based on a riskassessment tool developed in early February [11]. Note that Australian citizens and residents (and their dependants) were exempt from travel restrictions, but upon returning to Australia were required to quarantine for 14 days from the date of arrival. A full timeline of social distancing and border measures is provided in Figure S5.

**Figure 2:**
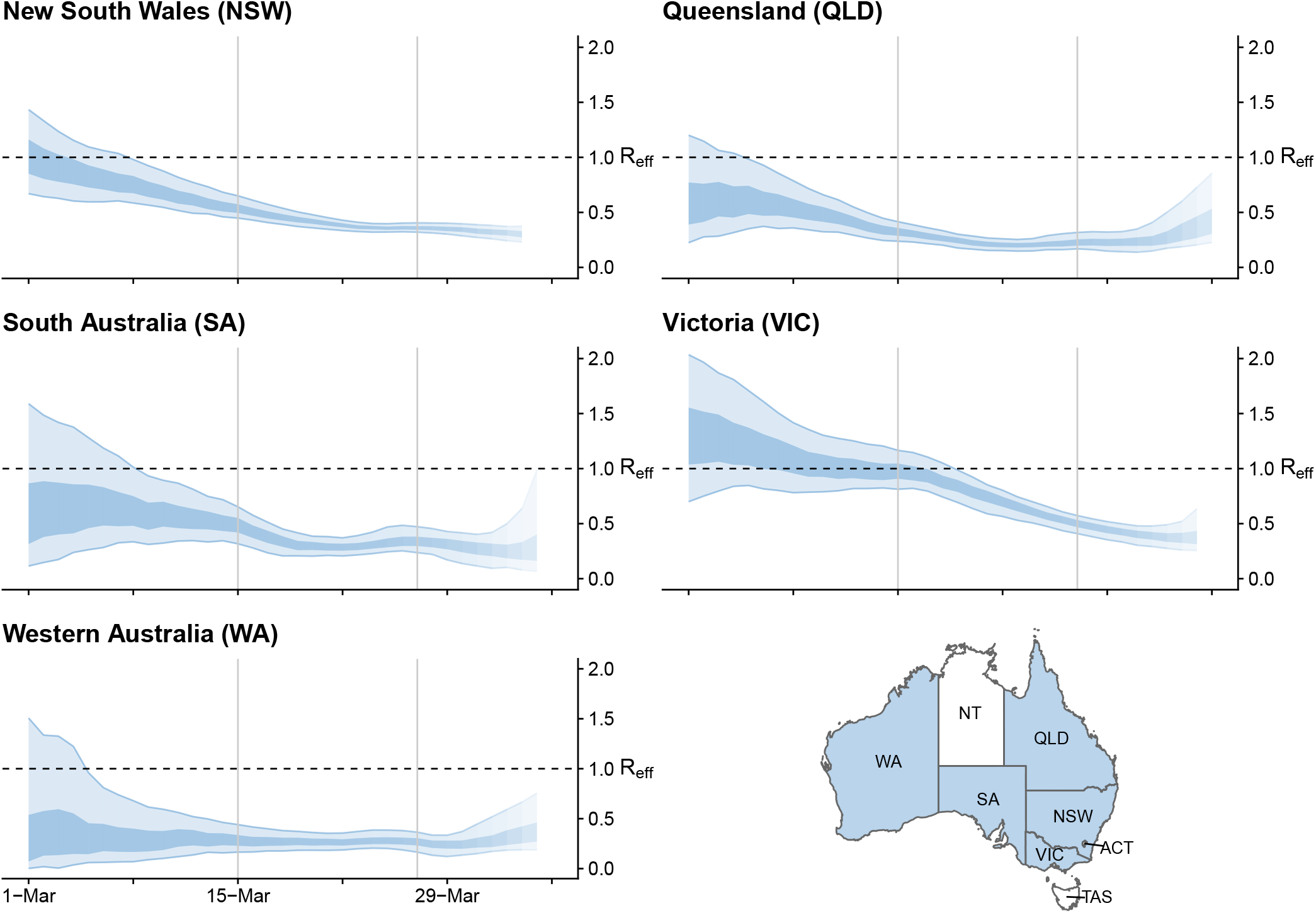
Time-varying estimate of the effective reproduction number (*R*_*eff*_) of COVID-19 by Australian state (light blue ribbon = 90% credible interval; dark blue ribbon = 50% credible interval) from 1 March to 5 April 2020, based on data up to and including 13 April 2020. Confidence in the estimated values is indicated by shading with reduced shading corresponding to reduced confidence. The horizontal dashed line indicates the target value of 1 for the effective reproduction number required for control. Not presented are the Australian Capital Territory (ACT), Northern Territory (NT) and Tasmania (TAS), as these states/territories had insufficient local transmission.

**Figure 3:**
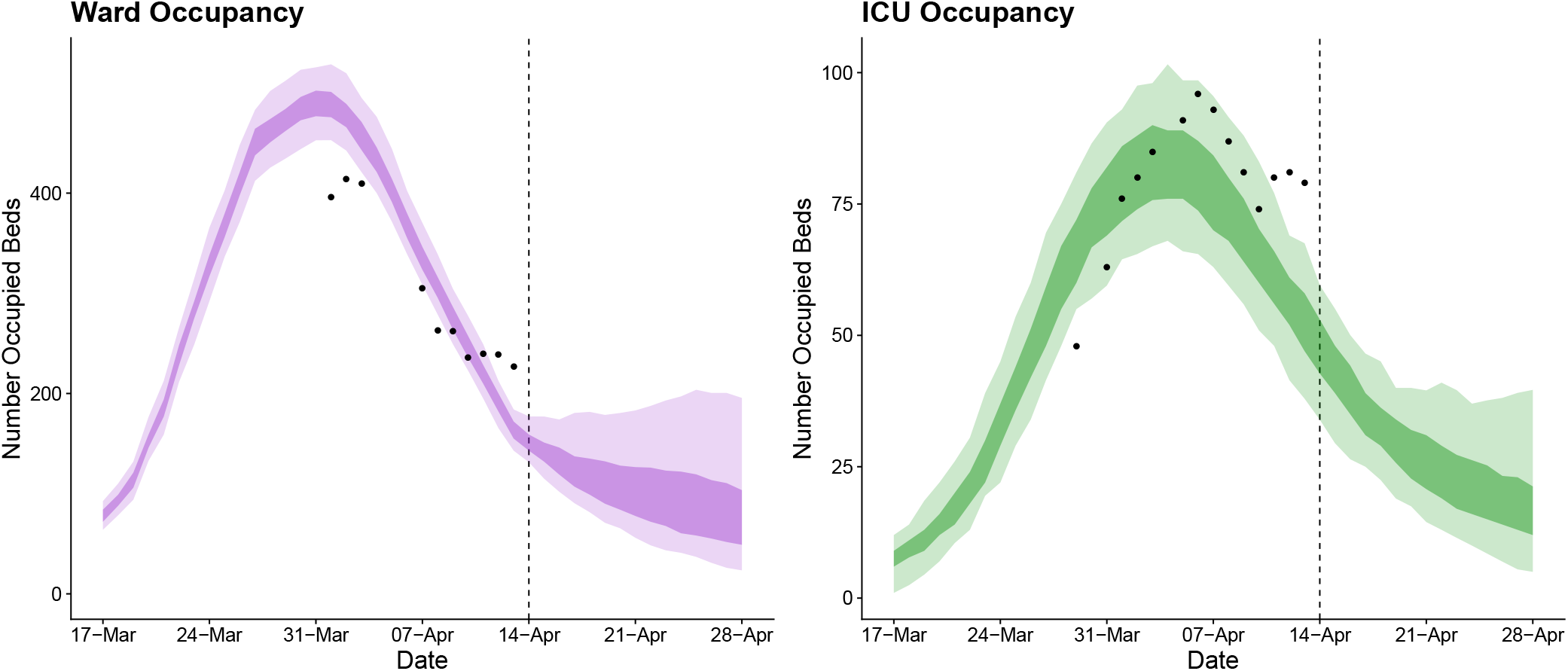
Forecasted daily hospital ward (left) and intensive care unit (right) occupancy (dark ribbons = 50% confidence intervals; light ribbons = 95% confidence intervals) from 17 March to 28 April. Occupancy = the number of beds occupied by COVID-19 patients on a given day. Black dots indicate the reported ward and ICU occupancy extracted from the Australian national COVID-19 database. Australian health system ward and ICU bed capacities are estimated to be over 25,000 and 1,100, respectively, under the assumption that 50% of total capacity could possibly be dedicated to COVID-19 patients [30]. The forecasted daily case counts are shown in Figure S6.

Australia took an early and precautionary approach to COVID-19. On 1 February, when China was the only country reporting uncontained transmission, Australian authorities restricted all travel from mainland China to Australia, in order to reduce the risk of importation of the virus. Only Australian citizens and residents (and their dependants) were permitted to travel from China to Australia. These individuals were advised to self-quarantine for 14 days from their date of arrival [10]. Further border measures, including enhanced testing and provision of additional advice, were placed on arrivals from other countries, based on a risk-assessment tool developed in early February [11].

The day before Australia imposed these restrictions (January 31), 9,720 cases of COVID-19 had been reported in mainland China [12]. Australia had so far detected and managed 9 imported cases, all with recent travel history from or a direct epidemiological link to Wuhan [13]. Before the restrictions, Australia was expecting to receive approximately 200,000 air passengers from mainland China during February 2020 [14]. Travel numbers fell dramatically following the imposed travel restrictions.

These restrictions were not intended (and highly unlikely [15]) to prevent the ultimate importation of COVID-19 into Australia. Their purpose was to delay the establishment of an epidemic, buying valuable time for health authorities to plan and prepare.

During the month of February, with extensive testing and case targeted interventions in place (case isolation and contact quarantine), Australia detected and managed only 12 cases. Meanwhile, globally, the geographic extent of transmission and daily counts of confirmed cases and deaths continued to increase drastically [16]. In early March, Australia extended travel restrictions to a number of countries with large uncontained outbreaks, namely Iran (as of 1 March) [17], South Korea (as of 5 March) [18] and Italy (as of 11 March) [19].

Despite these measures, the daily case counts rose sharply in Australia during the first half of March. While the vast majority of these cases were connected to travellers returning to Australia from overseas, localised community transmission had been reported in areas of Sydney and Melbourne. Crude plots of the cumulative number of cases by country showed Australia on an early trajectory similar to the outbreaks experienced in China, Europe and the United States, where health systems had become or were becoming overwhelmed [20].

From 16 March, the Australian Government progressively implemented a range of social distancing measures in order to reduce and prevent further community transmission [21]. The day before, authorities had imposed a self-quarantine requirement on all international arrivals [22]. On 19 March, Australia closed its borders to all non-citizens and non-residents [23], and on March 27, moved to a policy of mandatory quarantine for any returning citizens and residents [24]. By 29 March, social distancing measures had been escalated to the extent that all Australians were strongly advised to leave their homes only for limited essential activities and public gatherings were limited to two people [25].

By late March, daily counts of new cases appeared to be declining, suggesting that these measures had successfully reduced transmission.

Quantifying changes in the rate of spread of infection over the course of an epidemic is critical for monitoring the collective impact of public health interventions and forecasting the short-term clinical burden. A key indicator of transmission in context is the effective reproduction number (*R*_*eff*_) — the average number of secondary infections caused by an infected individual in the presence of public health interventions and for which no assumption of 100% susceptibility is made. If control efforts are able to bring *R*_*eff*_ below 1, then on average there will be a decline in the number of new cases reported. The decline will become apparent after a delay of approximately one incubation period plus time to case detection and reporting following implementation of the control measure (*i*.*e*., at least two weeks).

Using case counts from the Australian national COVID-19 database, we estimated *R*_*eff*_ over time for each Australian state from 24 February to 5 April 2020 (Figure 2). We used a statistical method that estimates time-varying *R*_*eff*_ by using an optimally selected moving average window to smooth the curve and reduce the impact of localised clusters and outbreaks that may cause large fluctuations [26]. Importantly, the method accounts for time delays between illness onset and case notification. Incorporation of this lag is critical for accurate interpretation of the most recent data in the analysis, to be sure that an observed drop in the number of reported cases reflects an actual drop in case numbers.

Results show that *R*_*eff*_ has likely been below 1 in each Australian state since early-to-mid March. These estimates are geographically averaged results over large areas and it is possible that *R*_*eff*_ was and remains much higher than 1 in a number of localised settings (see Figure 2).

The estimated time-varying *R*_*eff*_ value is based on cases that have been identified as a result of local transmission, whereas imported cases only contribute to the force of infection [27]. Imported and locally acquired cases were assumed to be equally infectious. The method for estimating *R*_*eff*_ is sensitive to this assumption. Hence, we performed a sensitivity analysis to assess the impact of stepwise reductions in the infectiousness of imported cases on *R*_*eff*_ as a result of quarantine measures implemented over time (see Figures S2, S3, and S4). The sensitivity analyses suggest that *R*_*eff*_ may well have dropped below 1 later than shown in Figure 2.

Next we used our estimates of time-varying *R*_*eff*_ to forecast the short-term clinical burden in Australia. Estimates were input into a mathematical model of disease dynamics that was extended to account for imported cases. A sequential Monte Carlo method was used to infer the model parameters and appropriately capture the uncertainty [28], conditional on each of a number of sampled *R*_*eff*_ trajectories up to 5 April, from which point they were assumed to be constant. The model was subsequently projected forward from April 14 to April 28, to forecast the number of reported cases, assuming a symptomatic detection probability of 80%, within the range estimated by [29].

The number of new daily hospitalisations and ICU admissions were estimated from recently observed and forecast case counts. Specifically, the age distribution of projected cases, and age-specific probabilities of hospitalisation and ICU admission, were extracted from Australian age-specific data on confirmed cases, assuming that this distribution would remain unchanged (see Table S2). In order to calculate the number of occupied ward/ICU beds per day, length-of-stay in a ward bed and ICU bed were assumed to be Gamma distributed with means (SD) of 11 (3.42) days and 14 (5.22) days, respectively. Our results indicate that with the current public health interventions in place Australia’s hospital ward and ICU occupancy will remain well below capacity thresholds over the next two weeks.

Our analysis suggests that Australia’s combined strategy of early, targeted management of the risk of importation, case targeted interventions, and broad-scale social distancing measures applied prior to the onset of detected widespread community transmission has substantially mitigated the first wave of COVID-19. More detailed analyses are required to assess the relative impact of specific response measures, and this information will be crucial for the next phase of response planning.

We further anticipate that the Australian health care system is well positioned to manage projected COVID-19 case loads over the next two weeks. Ongoing situational assessment and monitoring of forecasted hospital and ICU demand will be essential for managing possible future relaxation of broad-scale community interventions. Vigilance for localised increases in epidemic activity and in particular for outbreaks in vulnerable populations such as residential aged care facilities, where a high proportion of cases are likely to be severe, must be maintained.

While the symptomatic case detection rate is estimated to be very high in Australia (between 77 and 100% [29]), one largely unknown factor at present is the number of asymptomatic, mild and undiagnosed infections. Even if this number is high, the Australian population would still be largely susceptible to infection. Accordingly, complete relaxation of the measures currently in place would see a rapid resurgence in epidemic activity. This problem is not unique to Australia. Many countries with intensive social distancing measures in place are starting to grapple with their options and time frames for a gradual return to relative normalcy [31].

There are difficult decisions ahead for governments, and for now Australia is one of the few countries fortunate enough to be able to plan the next steps from a position of relative calm as opposed to crisis.

## Data Availability

Datasets analysed and generated during this study are included in the supplementary materials. For estimates of the time-varying effective reproduction number (Figure 2), the complete line listed data within the Australian national COVID-19 database are not publicly available. However, we provide the cases per day by notification date and state (as shown in Figures 1 and S1) which, when supplemented with the estimated distribution of the delay from symptom onset to notification (samples from this distribution are provided as a data file), analyses of the time-varying effective reproduction number can be performed.

## Acknowledgements

This study represents surveillance data reported through the Communicable Diseases Network Australia (CDNA) as part of the nationally coordinated response to COVID-19. We thank public health staff from incident emergency operations centres in state and territory health departments, and the Australian Government Department of Health, along with state and territory public health laboratories. We thank members of CDNA for their feedback and perspectives on the study results. We thank Dr Jonathan Tuke for helping to assemble Australian national and state announcements of COVID-19 response measures.

## Code availability

Analysis code is included in the supplementary materials.

## Methods

### Estimating the time-varying effective reproduction number

#### Overview

The method used to estimate *R*_*eff*_ is described in [32], as implemented in the R package, EpiNow [33]. This method is currently in development by the Centre for the Mathematical Modelling of Infectious Diseases at the London School of Hygiene & Tropical Medicine [26]. Full details of their statistical analysis and code base is available via their website (https://epiforecasts.io/covid/).

We provide a brief overview of the method below, focusing on how the analysis was adapted to the Australian context.

#### Data

We used line-lists of reported cases for each Australian state/territory extracted from the national COVID-19 database. The line-lists contain the date when the individual first exhibited symptoms, date when the case notification was received by the jurisdictional health department and where the infection was acquired (*i*.*e*., overseas or locally).

#### Reporting delays and under-reporting

A *pre-hoc* statistical analysis was conducted in order to estimate a distribution of the reporting delays from the line-lists of cases, using the code base provided by [26]. The estimated reporting delay is assumed to remain constant over time. These reporting delays are used to: i) infer the time of symptom onset for those without this information, and; ii) infer how many cases in recent days are yet to be recorded. Adjusting for reporting delays is critical for inferring when a drop in observed cases reflects a true drop in cases.

Trends identified using this approach are robust to under-reporting, assuming that it is constant. However, absolute values of *R*_*eff*_ may be biased by reporting rates. Pronounced changes in reporting rates may also impact the trends identified. However, evidence suggests that Australia’s symptomatic case ascertainment rate is very high (between 77 and 100%) and that this rate has been relatively stable over time [29].

#### Estimating the effective reproduction number over time

Briefly, the *R*_*eff*_ was estimated for each day from 24 February 2020 up to 5 April 2020 using line list data – date of symptom onset, date of report, and import status – for each state. The method assumes that the serial interval (*i*.*e*., time between symptom onset for an index and secondary case) is uncertain, with a mean of 4.7 days (95% CrI: 3.7, 6.0) and a standard deviation of 2.9 days (95% CrI: 1.9, 4.9), as estimated from early outbreak data in Wuhan, China [34]. Combining the incidence over time with the uncertain distribution of serial intervals allows us to estimate *R*_*eff*_ over time.

A prior distribution was specified for *R*_*eff*_, with mean 2.6 (informed by [35]) and a broad standard deviation of 2 so as to allow for a range of *R*_*eff*_ values. Finally, *R*_*eff*_ is estimated using an optimally selected moving average window in order to smooth the curve and reduce the impact of localised events (*e*.*g*., local outbreaks) causing large variations.

Note that up to 20% of reported cases in the Australian national COVID-19 database do not have a reported import status (see Figure 1). Conservatively, we assumed that all cases with an unknown or unconfirmed source of acquisition were locally acquired.

#### Accounting for imported cases

A large proportion of cases reported in Australia from January until now were imported from overseas. It is critical to account for two distinct populations in the case notification data — imported and locally acquired — in order to perform robust analyses of transmission in the early stages of this outbreak. The estimated time-varying *R*_*eff*_ value is based on cases that have been identified as a result of local transmission, whereas imported cases contribute to transmission only [27].

Specifically, the method assumes that local and imported cases contribute equally to transmission. The results under this assumption are presented in Figure 2. However, given quarantine measures put in place at different times (Figure S5), it is likely that imported cases contributed relatively less to transmission than locally acquired cases. We explored this via a sensitivity analysis. Prior to 15 March, returning Australian residents and citizens (and their dependents) from mainland China were advised to self-quarantine. Note that further border measures were implemented during this period, including enhanced testing and provision of advice on arrivals from selected countries based on a risk assessment tool developed in early February [11]. On 15 March, Australian authorities imposed a *self-quarantine* requirement on all international arrivals, and from 27 March, moved to a *mandatory* quarantine policy for all international arrivals. Hence, we assumed that prior to 15 March, 10%, 20% and 50% of imported cases did not contribute to transmission (Figures S2, S3 and S4, respectively). Between 15 and 27 March, we assumed that 50%, 50% and 80% of imported cases did not contribute to transmission. From 27 March, we assumed that 99% of imported cases did not contribute to transmission in each scenario.

### Forecasting short-term ward and ICU bed occupancy

We used the estimates of time-varying *R*_*eff*_ to forecast the national short-term ward/ICU occupancy due to COVID-19 patients.

#### Forecasting case counts

The forecasting method combines an SEEIIR (susceptible-exposed-infectious-recovered) population model of infection with daily COVID-19 case notification counts, through the use of a bootstrap particle filter. The daily case counts by date of diagnosis were modelled using a negative binomial distribution with a fixed dispersion parameter *k*, and the expected number of cases was proportional to the daily incidence of symptomatic infections in the SEEIIR model; this proportion was characterised by the observation probability. Natural disease history parameters were sampled from narrow uniform priors, based on values reported in the literature for COVID-19, and each particle was associated with an *R*_*eff*_ trajectory that was drawn from the state/territory *R*_*eff*_ trajectories in Figure 2 up to 5 April, from which point they are assumed to be constant. The model was subsequently projected forward from April 14 to April 28, to forecast the number of reported cases, assuming a detection probability of 80% (informed by [29]).

In order to account for imported cases, we used daily counts of imported cases to construct a time-series of the expected daily importation rate and, assuming that such cases were identified one week after initial exposure, introduced exposure events into each particle trajectory by adding an extra term to the force of infection equation.

Model equations below describe the flow of individuals in the population from the susceptible class (*S*), through two exposed classes (*E*_1_, *E*_2_), two infectious classes (*I*_1_, *I*_2_) and finally into a removed class (*R*). Two exposed and infectious classes are chosen such that the duration of time in the exposed or infectious period has an Erlang distribution. The corresponding parameters are given in Table S1.

Model equations:

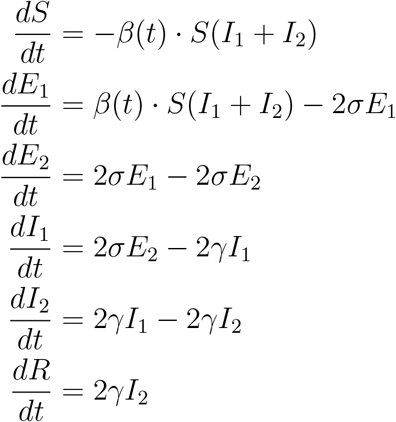

With initial conditions:

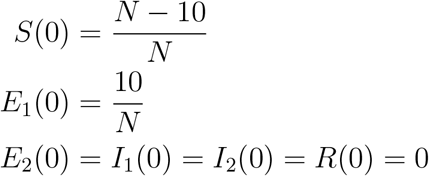

Observation model:

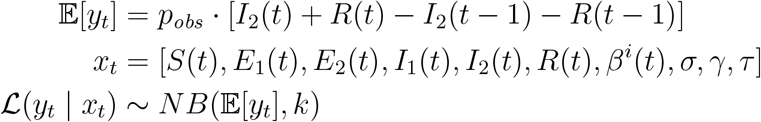

With time-varying transmission rate:

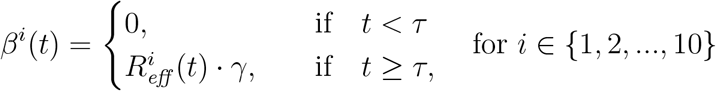

**Table S1:** SEEIIR forecasting model parameters.

| Parameter | Definition | Value/Prior distribution |
| --- | --- | --- |
| $\sigma$ | Inverse of the mean incubation period | $U(4^{-1}, 3^{-1})$ |
| $\gamma$ | Inverse of the mean infectious period | $U(10^{-1}, 9^{-1})$ |
| $\tau$ | Time of first exposure | $U(0, 28)$ |
| $p_{obs}$ | Probability of observing a case | 0.8 |
| $k$ | Dispersion parameter on Negative-Binomial observation model | 100 |

#### Forecasting ward and ICU bed occupancy from observed and projected case counts

The number of new daily hospitalisations and ICU admissions were estimated from recently observed and forecasted case counts by:

1. Estimating the age distribution of projected case counts using data from the national COVID-19 database on the age-specific proportion of confirmed cases;
2. Estimating the age-specific hospitalisation and ICU admission rates using data from the national COVID-19 database. We assumed that all hospitalisations and ICU admissions were either recorded or were missing at random (31% and 58% of cases had no information recorded under hospitalisation or ICU status, respectively);
3. Randomly drawing the number of hospitalisations/ICU admissions in each age-group (for both the observed and projected case counts) from a binomial distribution with number of trials given by the expected number of cases in each age group (from 1), and probability given by the observed proportion of hospitalisations/ICU admissions by age group (from 2).

Finally, in order to calculate the number of occupied ward/ICU beds per day, length-of-stay in a ward bed and ICU bed were assumed to be Gamma distributed with means (SD) of 11 (3.42) days and 14 (5.22) days, respectively. We assumed ICU admissions required a ward bed prior to, and following, ICU stay for a Poisson distributed number of days with mean 2.5. Relevant Australian data were not available to parameterise a model that captures the dynamics of patient flow within the hospital system in more detail. This model provides a useful indication of hospital bed occupancy based on limited available data and may be updated as more specific data (*e*.*g*., on COVID-19 patient length-of-stay) becomes available.

**Table S2:**
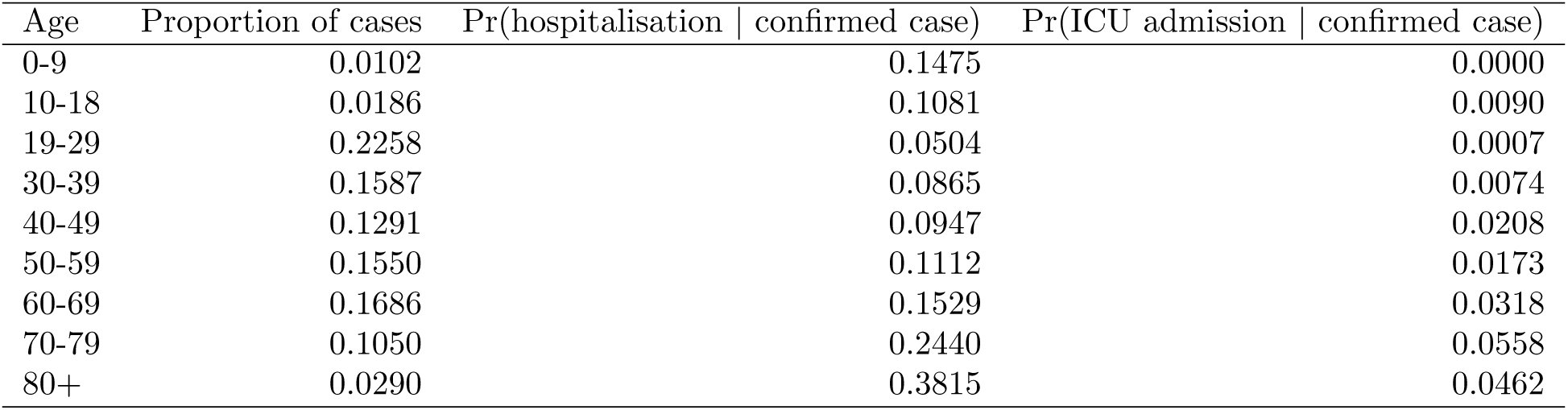
Age-specific proportions of confirmed cases extracted from the Australian national COVID-19 database and age-specific estimates of the probability of hospitalisation and ICU admission for confirmed cases.

**Figure S1:**
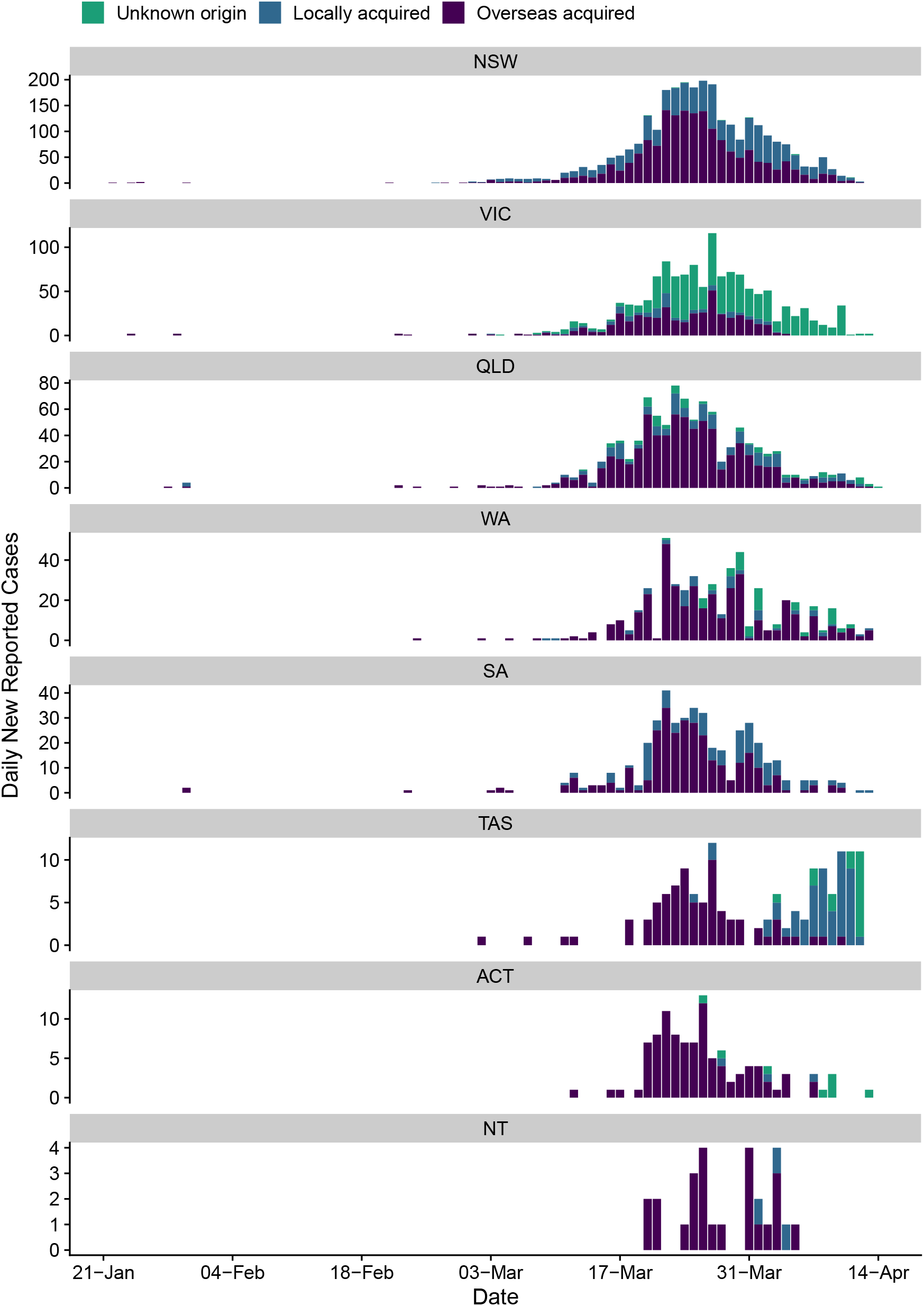
Time series of new daily confirmed cases of COVID-19 in each Australian state/territory by import status (purple = overseas acquired, blue = locally acquired, green = unknown origin) from 22 January 2020 (first case detected) to 13 April 2020.

**Figure S2:**
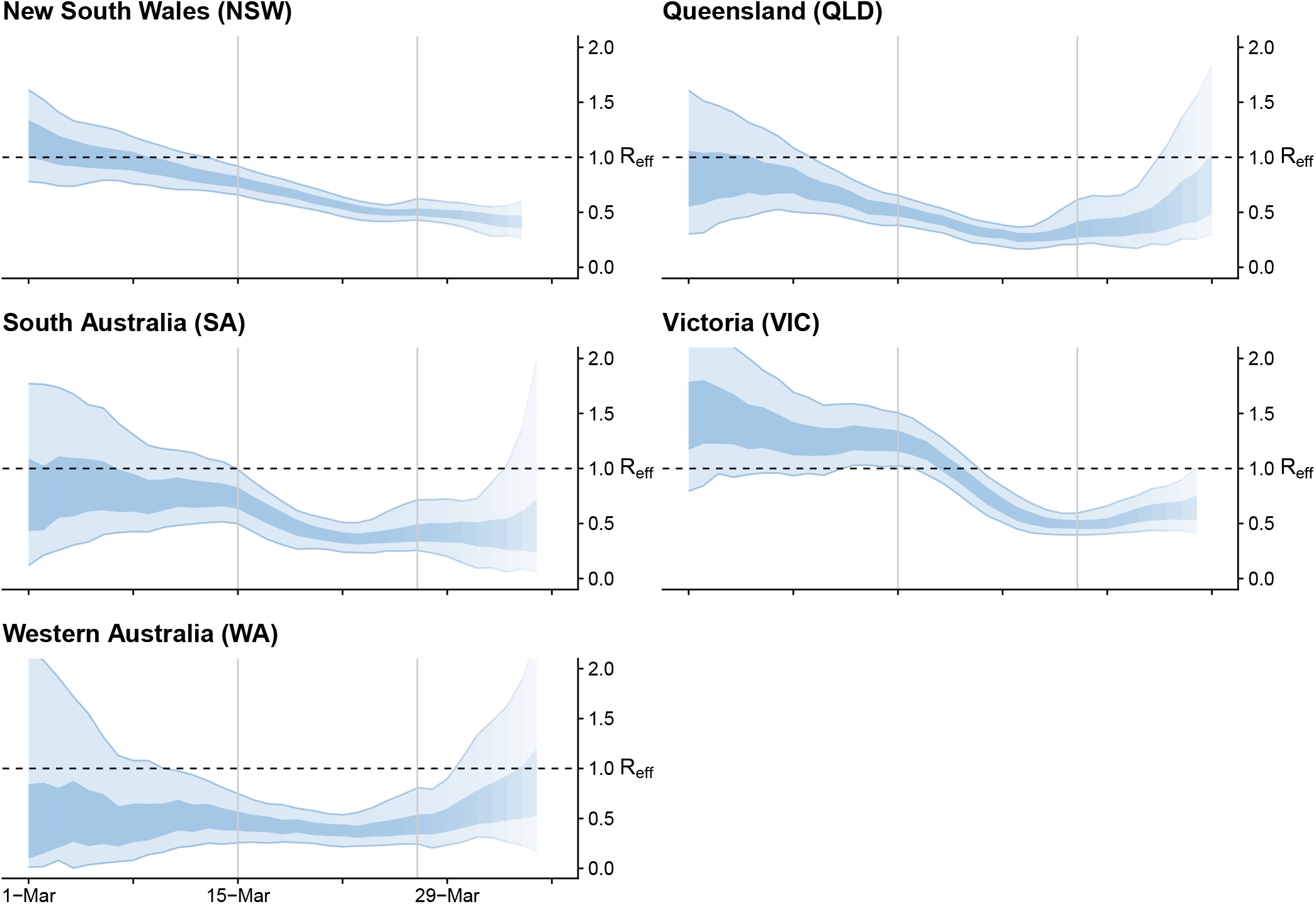
Sensitivity analysis 1 of 3. Time-varying estimate of the effective reproduction number (*R*_*eff*_) of COVID-19 by Australian state (light blue ribbon = 90% credible interval; dark blue ribbon = 50% credible interval) from 1 March to 5 April 2020, based on data up to and including 13 April 2020. Confidence in the estimated values is indicated by shading, with reduced shading corresponding to reduced confidence. The horizontal dashed line indicates the target value of 1 for the effective reproduction number required for control. Results are produced assuming stepwise changes in the relative infectiousness of locally acquired to imported cases according to quarantine requirements for returning travellers introduced on 15 and 27 March (indicated by vertical grey lines). We assumed 10%, 50%, and 99% of imported cases did not contribute to transmission prior to 15 March, between 15 and 27 March (inclusive), and after 27 March, respectively.

**Figure S3:**
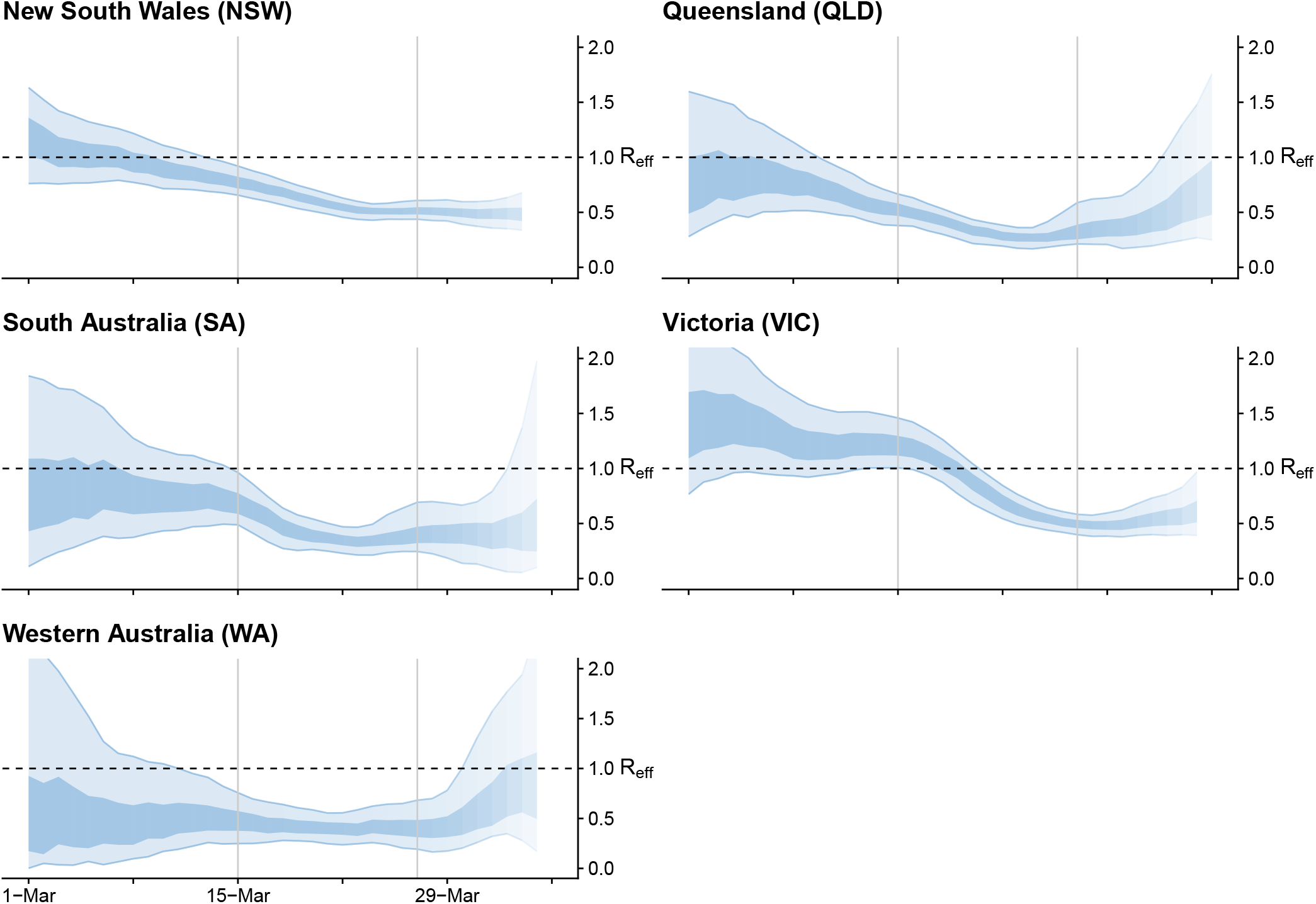
Sensitivity analysis 2 of 3. Time-varying estimate of the effective reproduction number (*R*_*eff*_) of COVID-19 by Australian state (light blue ribbon = 90% credible interval; dark blue ribbon = 50% credible interval) from 1 March to 5 April 2020, based on data up to and including 13 April 2020. Confidence in the estimated values is indicated by shading, with reduced shading corresponding to reduced confidence. The horizontal dashed line indicates the target value of 1 for the effective reproduction number required for control. Results are produced assuming stepwise changes in the relative infectiousness of locally acquired to imported cases according to quarantine requirements for returning travellers introduced on 15 and 27 March (indicated by vertical grey lines). We assumed 20%, 50%, and 99% of imported cases did not contribute to transmission prior to 15 March, between 15 and 27 March (inclusive), and after 27 March, respectively.

**Figure S4:**
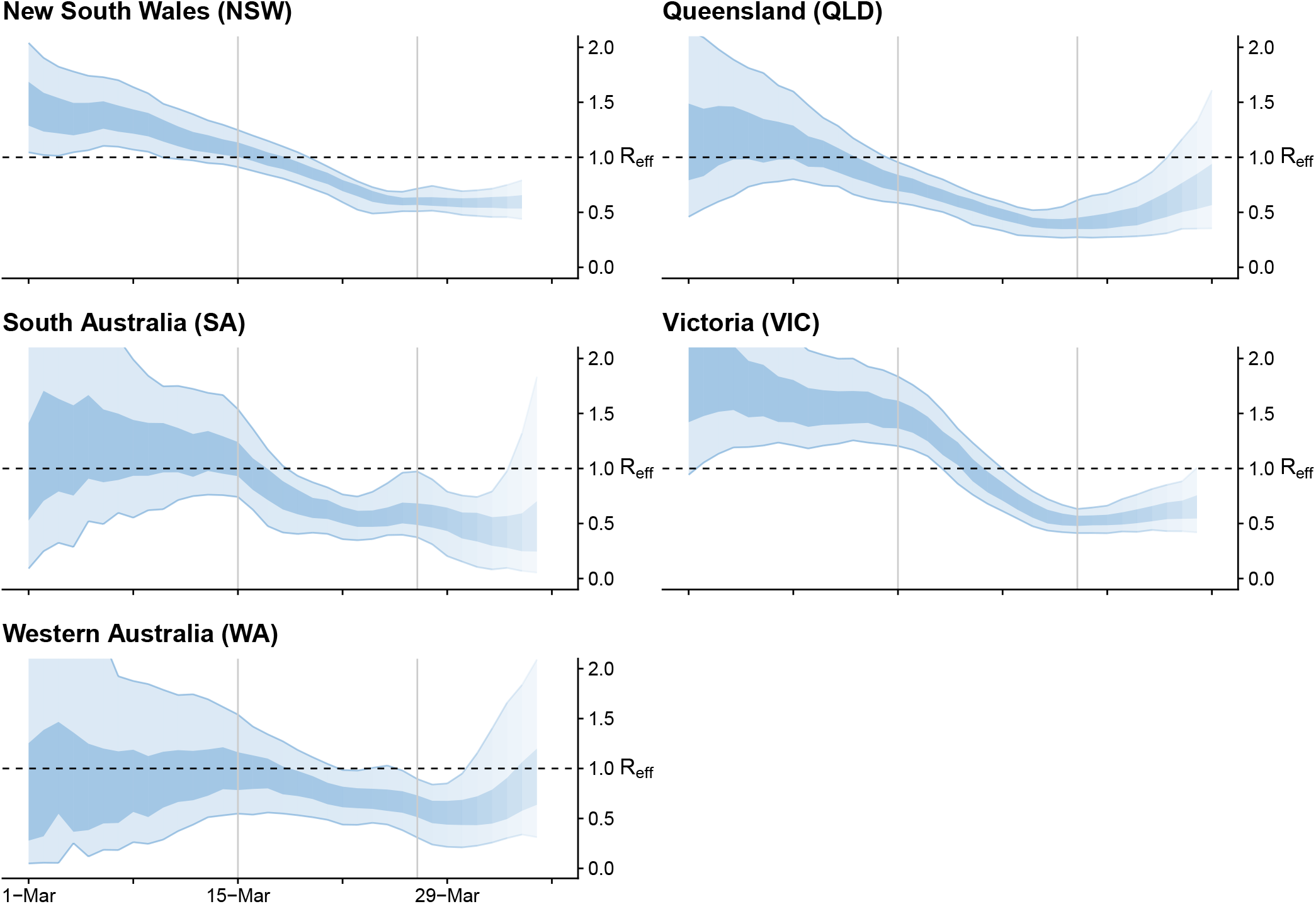
Sensitivity analysis 3 of 3. Time-varying estimate of the effective reproduction number (*R*_*eff*_) of COVID-19 by Australian state (light blue ribbon = 90% credible interval; dark blue ribbon = 50% credible interval) from 1 March to 5 April 2020, based on data up to and including 13 April 2020. Confidence in the estimated values is indicated by shading, with reduced shading corresponding to reduced confidence. The horizontal dashed line indicates the target value of 1 for the effective reproduction number required for control. Results are produced assuming stepwise changes in the relative infectiousness of locally acquired to imported cases according to quarantine requirements for returning travellers introduced on 15 and 27 March (indicated by vertical grey lines). We assumed 50%, 80%, and 99% of imported cases did not contribute to transmission prior to 15 March, between 15 and 27 March (inclusive), and after 27 March, respectively.

**Figure S5:**
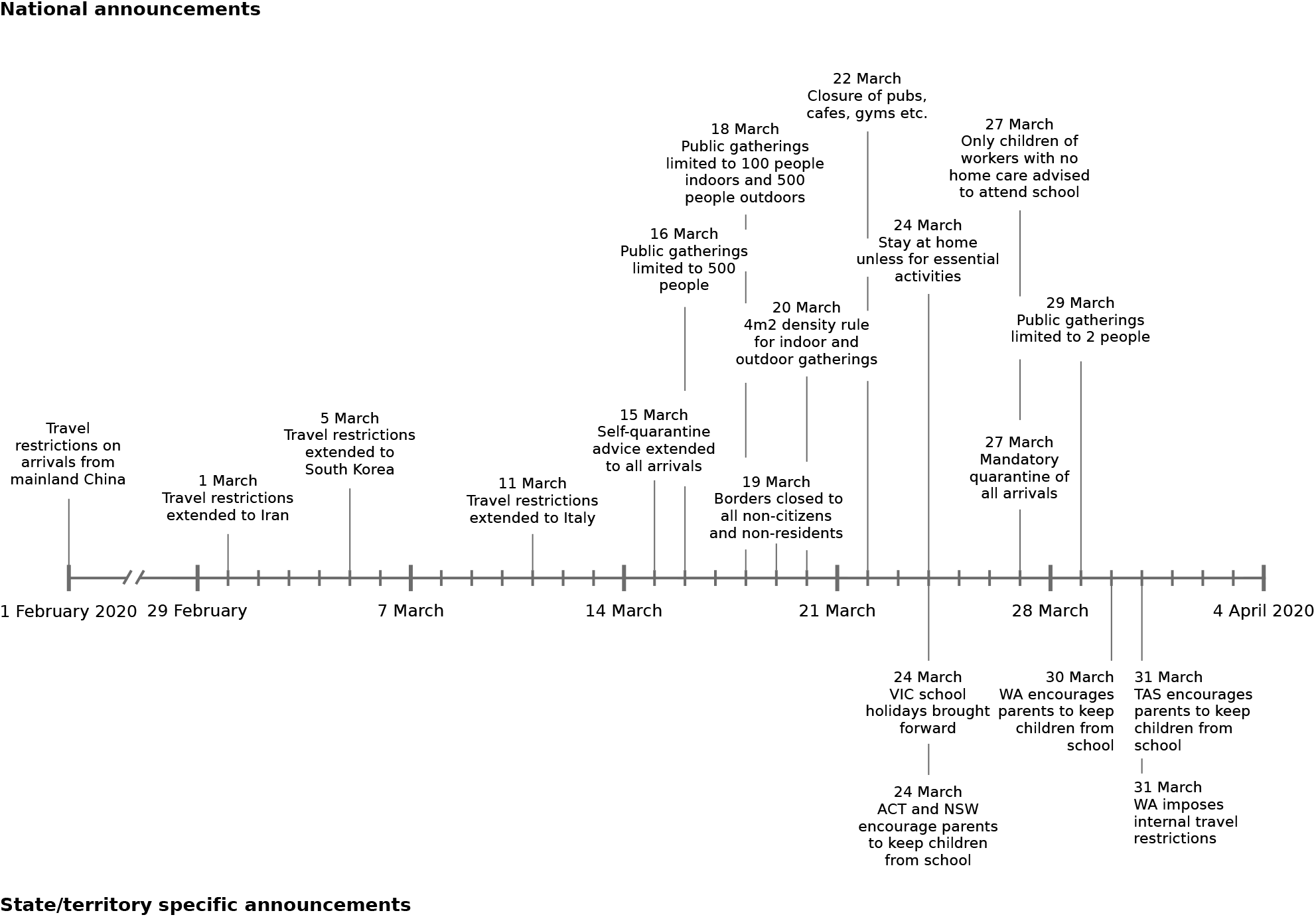
Timeline of border and social distancing measures implemented in Australia up to 4 April 2020. These measures were in addition to case targeted interventions, specifically case isolation and quarantine of their contacts. Note 1: Between 1 February and 15 March, further border measures were introduced, including enhanced testing and provision of advice on arrivals from other selected countries, based on a risk-assessment tool developed in early February [11]. Note 2: Australian citizens and residents (and their dependants) were exempt from travel restrictions but upon returning to Australia were required to quarantine for 14 days from the date of arrival.

**Figure S6:**
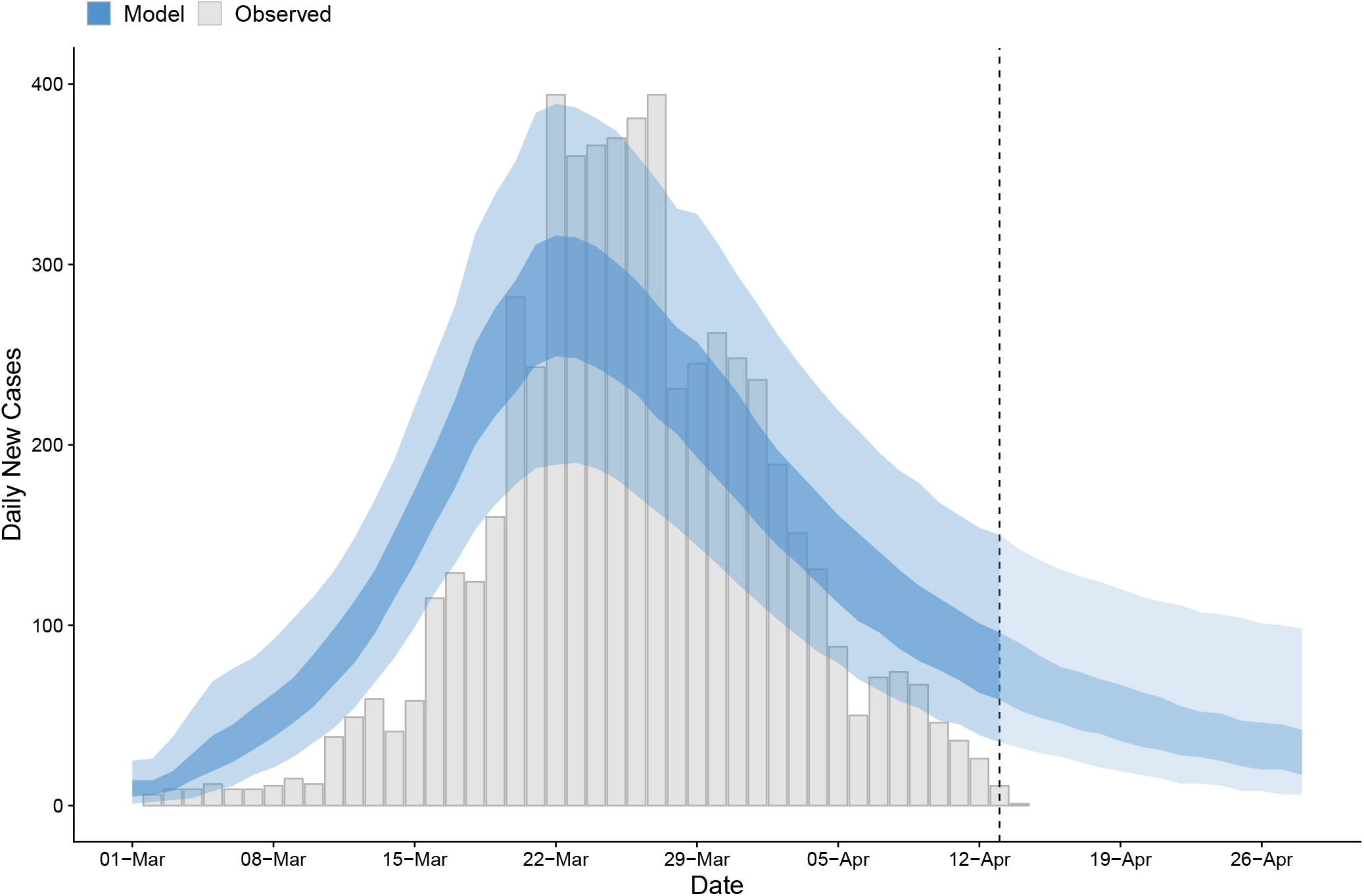
Time series of new daily confirmed cases of COVID-19 in Australia from 1 March to 13 April 2020 (grey bars) overlaid by daily case counts estimated from the forecasting model up to April 13 and projected forward from 14 to 28 April inclusive. Inner shading = 50% confidence intervals. Outer shading = 95% confidence intervals. Note that forecasting model estimates prior to 13 April — the last recorded data point at the time of analysis (indicated by the dashed grey line) — use data up to and including the previous day.

